# The socio-economic determinants of the coronavirus disease (COVID-19) pandemic

**DOI:** 10.1101/2020.04.15.20066068

**Authors:** Viktor Stojkoski, Zoran Utkovski, Petar Jolakoski, Dragan Tevdovski, Ljupco Kocarev

## Abstract

The magnitude of the coronavirus disease (COVID-19) pandemic has an enormous impact on the social life and the economic activities in almost every country in the world. Besides the biological and epidemiological factors, a multitude of social and economic criteria also govern the extent of the coronavirus disease spread in the population. Consequently, there is an active debate regarding the critical socio-economic determinants that contribute to the resulting pandemic. In this paper, we contribute towards the resolution of the debate by leveraging Bayesian model averaging techniques and country level data to investigate the potential of 35 determinants, describing a diverse set of socio-economic characteristics, in explaining the coronavirus pandemic outcome.

## 1 Introduction

The coronavirus pandemic began as a simple outbreak in December 2019 in Wuhan, China. However, it quickly propagated to other countries and became a primary global threat. It seems that most countries were not prepared for this pandemic. As a consequence, hospitals were over-crowded with patients and death rates due to the disease skyrocketed. In particular, as of the time of this writing (11th April 2020), there have been over 1.5 million cases and over 100 thousand deaths worldwide as a cause of the coronavirus induced disease, COVID-19^1^.

In order to reduce the impact of the disease spread, most governments implemented social distancing restrictions such as closure of schools, airports, borders, restaurants and shopping malls [1]. In the most severe cases there were even lockdowns – all citizens were prohibited from leaving their homes. This subsequently lead to a major economic downturn: stock markets plummeted, international trade slowed down, businesses went bankrupt and people were left unemployed. While in some countries the implemented restrictions had a significant impact on reducing the expected shock from the coronavirus, the extent of the disease spread in the population greatly varied from one economy to another, as illustrated in Fig 1.

**Figure 1:**
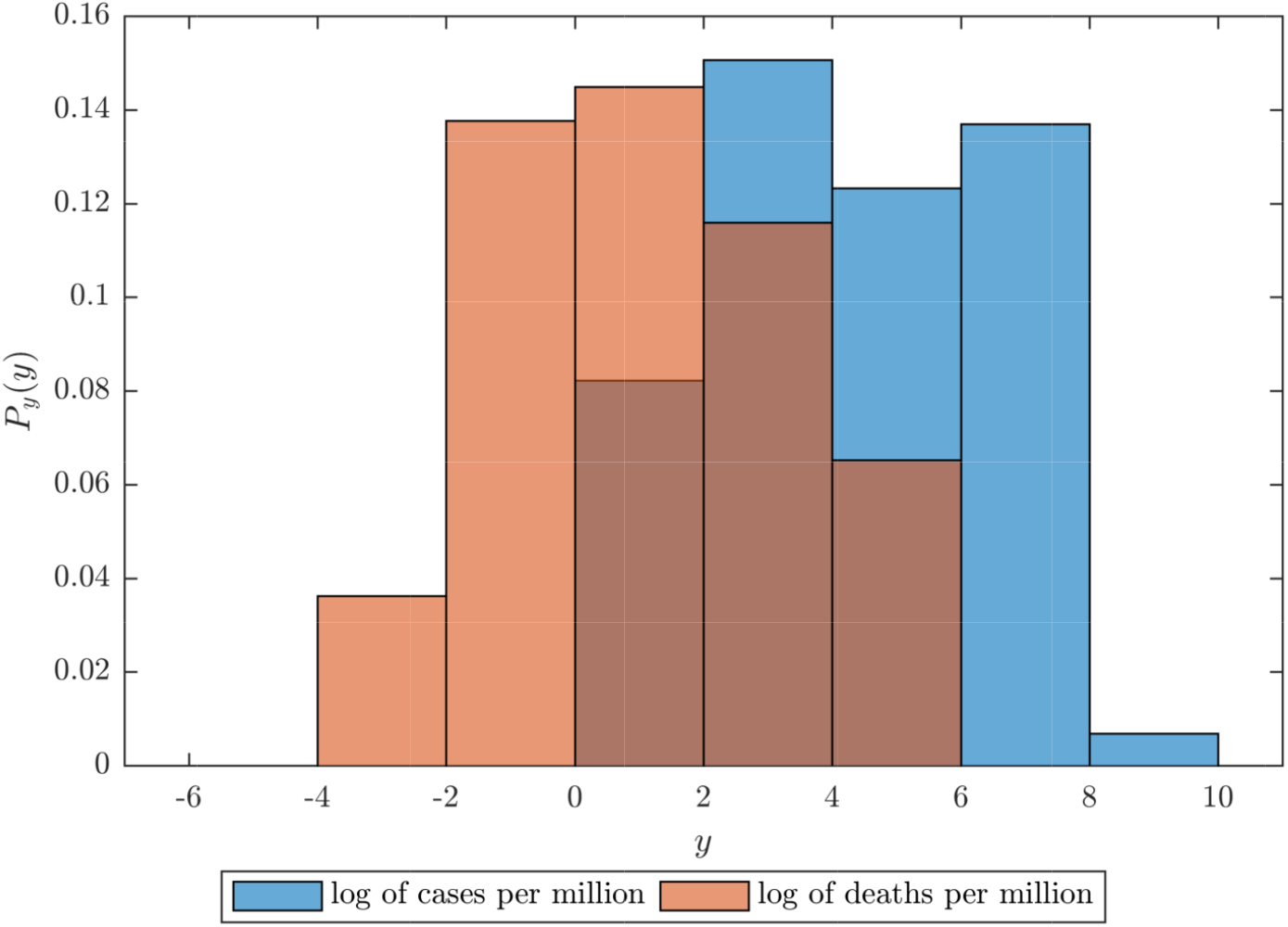
Histogram based on probability density estimation for the cases and deaths per million population using country level data. The x-axis describes the observed value, whereas the y-axis is the estimated probability density. Data taken on 11th April 2020.

A multitude of social and economic criteria have been attributed as potential determinants for the observed variety in the coronavirus outcome. Some experts say that the hardest hit countries also had an aging population [2, 3], or an underdeveloped healthcare system [4, 5]. Others emphasize the role of the natural environment [6, 7]. In addition, while the developments in most of the countries follow certain common patterns, several countries are notably outliers, both in the number of documented cases and in the disease outcome.Having in mind the ongoing debate, a comprehensive empirical study of the critical socio-economic determinants of the coronavirus pandemic would not only provide a glimpse on their potential impact, but would also offer a guidance for future policies that aim at preventing the emergence of epidemics.

Motivated by this observation, here we perform a detailed statistical analysis on a large set of potential socio-economic determinants and explore their potential to explain the variety in the observed coronavirus cases/deaths among countries. To construct the set of potential determinants we conduct a thorough review of the literature describing the social and economic factors which contribute to the spread of an epidemic. We identify a total of 35 potential determinants that describe a diverse ensemble of social and economic factors, including: healthcare infrastructure, societal characteristics, economic performance, demographic structure etc. To investigate the performance of each variable in explaining the coronavirus outcome, we utilize the technique of Bayesian model averaging (BMA). BMA allows us to isolate the most important determinants by calculating the posterior probability that they truly regulate the process. At the same time, BMA provides estimates for their relative impact, while also accounting for the uncertainty in the selection of potential determinants [8–10].

Based on the current data, we observe patterns that suggest that there are only few determinants (factors) that have explanatory power for the coronavirus outcome. As we will discuss in more detail in the sequel, we observe that some of these factors are strongly related to the level of economic development and to the effect of population size in social interactions. However, we stress that at this point in time, our observations require a careful interpretation for several reasons, including the following: (i) the analysis is based on aggregate quantities, i.e. averages across geographic locations. As such, it does not include spatial inhomogeneity and hence can not capture (potentially) significant interactive local dynamics; (ii) the parameters governing the time evolution of the disease spread and pandemic outcome are themselves dynamic, and depend on the stage of the country’s epidemic and on the changing social response efforts. While being aware of these potential shortcomings of our formulation, in the absence of realistic models that adequately cover all relevant aspects, this study provides the first step towards a more comprehensive understanding of the socio-economic factors of the coronavirus pandemic. We expect that, with the availability of new data and the improved understanding of the dynamics of the coronavirus pandemic, some of these shortcomings will be overcome, yielding a more reliable interpretation of the results.

## 2 Results

### 2.1 Preliminaries

In a formal setting, both the log of registered COVID-19 cases and the log of COVID-19 deaths are a result of a disease spreading process [11, 12]. The extent to which a disease spreads within a population is uniquely determined by its reproduction number. This number describes the expected number of cases directly generated by one case in a population in which all individuals are susceptible to infection [13, 14]. Obviously, its magnitude depends on various natural characteristics of the disease, such as its infectivity or the duration of infectiousness [15], and the social distancing measures imposed by the government [1]. Also, it depends on a plethora on socio-economic factors that govern the behavioral interactions within a population [16, 17].

In general, we never observe the reproduction number, but instead its outcome, i.e. the number of cases/deaths. Hence, we can utilize the known properties of the reproduction number to derive a linear regression model *M*_*m*_ for the coronavirus outcome as

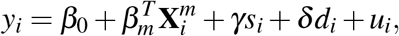

where for simplicity we denote both the log of registered COVID-19 cases per million population and the log of COVID-19 deaths per million population of country *i* as *y*_*i*_. We focus on registered quantities normalized on per capita basis for the dependent variable instead of raw values as a means to eliminate the bias in the outcome arising as a consequence of the discrepancies in the sizes of the studied countries. In the equation, 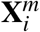 it is a *k*_*m*_ dimensional vector of socio-economic explanatory variables that determine the dependent variable, *β*_*m*_ is the vector describing their marginal contributions, *β*_0_ is the intercept of the regression, and *u*_*i*_ is the error term. The *s*_*i*_ term controls for the impact of social distancing measures of the countries, and *γ* is its coefficient. Finally, we also include the term *d*_*i*_, with *δ* capturing its marginal effect, that measures the duration of the pandemics within the economy. This allows us to control for the possibility that the countries are in a different state of the disease spreading process.

A central question which arises is the selection of the independent variables in *M*_*m*_. While the literature review offers a comprehensive overview of all potential determinants, in reality we are never certain of their credibility. In order to circumvent the problem of choosing a model and potentially ending up with a wrong selection, we resort to the technique of Bayesian Model Averaging (BMA). BMA leverages Bayesian statistics to account for model uncertainty by estimating each possible model, and thus evaluating the posterior distribution of each parameter value and probability that a particular model is the correct one [18].

### 2.2 Baseline model

The BMA method relies on the estimation of a baseline model *M*_0_ that is used for evaluating the performance of all other models. In our case, *M*_0_ is the model which encompasses only the effect of government social distancing measures and the duration of the pandemics in the country.

We measure the duration of pandemics in a country simply as the number of days since the first registered case, whereas in order to assess the effect of government restrictions we construct a stringency index. Mathematically, the index quantifies the average daily variation in government responses to the pandemic dynamics. As a measure for the daily variation we take the Oxford Covid-19 stringency index^2^. The Oxford Covid-19 stringency index is a composite measure that combines the daily effect of policies on school closures, reduction of internal movement, travel bans and other similar restrictions. For each country, we construct a weighted average of the index from all available data since their first registered coronavirus case up to the last date of data gathering. To emphasize the effect of policy restrictions implemented on an earlier date in calculating the average value, we put a larger weight on those dates. This is because earlier restrictions have obviously a bigger impact on the prevention of the spread of the virus. The procedure implemented to derive the average government stringency index is described in greater detail in Section S2.1 of the Supplementary Material (SM).

Fig 2 visualizes the results from the baseline model. We observe that the countries which had stringer policies also had less COVID-19 cases and deaths, as expected. In addition, the countries with longer duration of the crisis registered more cases and deaths per million population, though, the effect of this variable is negligible.

**Figure 2:**
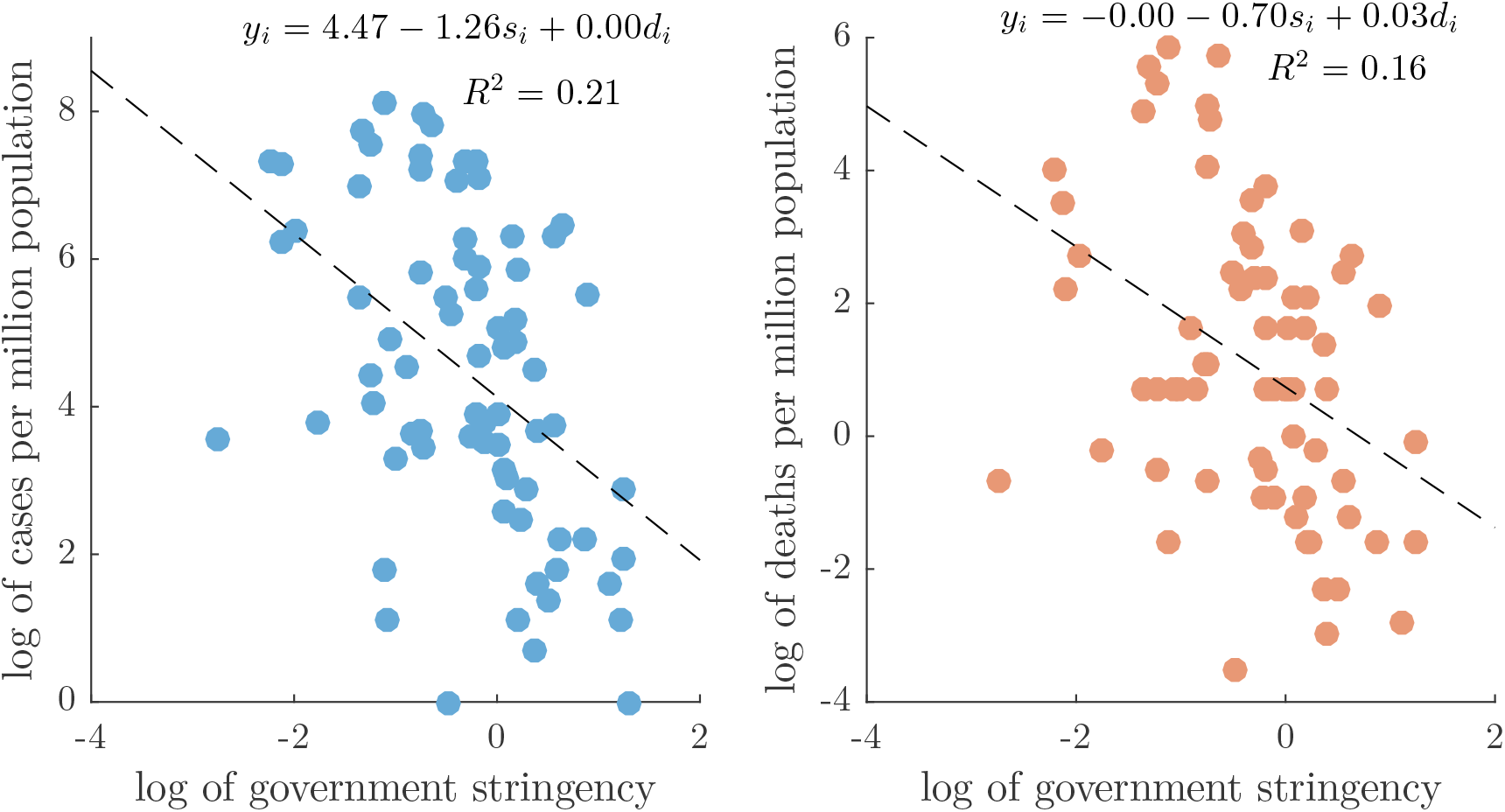
Explained variation in COVID-19 cases due to government stringency.

### 2.3 Socio-economic determinants

It is apparent that the baseline model can explain only a certain amount of the variations in registered covid cases/deaths. A fraction of the rest, we believe, can be attributed to various socio-economic determinants present within a society. To derive the set of potential determinants we conduct a comprehensive literature review. From the literature review we recognize a total of 35 potential socio-economic determinants, listed in Table 1. For a detailed description of the potential effect of the determinants we refer to the references given in the same table, and the references therein. In what follows, we only describe in short the potential determinants on the basis of the socio-economic characteristic they exhibit.

**Table 1:** List of Potential determinants of the COVID–19 pandemic.

| Determinant | Measure | Source | Refs. |
| --- | --- | --- | --- |
| **Healthcare Infrastructure** |  |  |  |
| Number of physicians | Physicians p.c. | WDI | [19–28] |
| Number of nurses | Nurses and midwives p.c. | WDI | [19–28] |
| Number of hospital beds | Hospital beds p.c. | WDI | [19–28] |
| Health coverage | UHC service coverage index | WDI | [19–28] |
| **National health statistics** |  |  |  |
| Birth Rate | Birth rate, crude p.c. | WDI | [29–32] |
| Death Rate | Death rate, crude p.c. | WDI | [29–32] |
| Life expectancy | Life expectancy at birth, (years) | WDI | [29–32] |
| Mortality from diabetes | Mortality rate from diabetes | WDI | [33, 34] |
| Mortality from communicable diseases | Mortality rate from com. diseases | WDI | [35] |
| Mortality from hygiene | Mortality rate from lack of hygiene | WDI | [36, 37] |
| **Economic performance** |  |  |  |
| Economic development | GDP p.c., PPP \$ | WDI | [38–41, 43, 44] |
| Labor market | Employment to population ratio (%) | WDI | [19, 38, 42] |
| Government spending | Gov. health spending p.c., PPP \$) | WDI | [26, 38–41] |
| Government debt | Government gross debt(% of GDP) | IMF | [38–41, 45, 46] |
| Income inequality | GINI index | WDI | [47–51] |
| Trade | Trade (% of GDP) | WDI | [19, 42, 52] |
| **Societal characteristics** |  |  |  |
| Media usage | % of population using internet | WDI | [19, 54–58] |
| Education | Human capital index | WDI | [29, 54–58] |
| Government effectiveness | Government effectiveness | WGI | [59, 60] |
| Rule of law | Rule of law | WGI | [59, 60] |
| Control of corruption | Control of corruption | WGI | [59, 60] |
| Religion | 60%+ catholic population | NM | [61–63] |
|  | 60%+ christian population | NM | [61–63] |
|  | 60%+ muslim population | NM | [61–63] |
| Household size | Avg. no. of persons in a household | UN | [17, 53, 64–66] |
| **Demographic structure** |  |  |  |
| Elderly population | Population age 65+ (% of total) | WDI | [67–69] |
| Young population | Population ages 0-14 (% of total) | WDI | [67–69] |
| Population size | Population, total | WDI | [70, 71] |
| Rural population | Rural population (% of total) | WDI | [70, 71] |
| Migration | Int. migrant stock (% of population) | WDI | [70, 71] |
| Population density | People per sq. km | WDI | [70, 71] |
| **Natural environment** |  |  |  |
| Sustainable development | Ecological Footprint (gha/person) | GFN | [6] |
| Air Pollution | Yearly avg P.M. 2.5 exposure | SGA | [7, 72, 73] |
| Air transport | Yearly passengers carried | WDI | [26] |
| International Tourism | Number of tourist arrivals | WDI | [26] |

#### Healthcare Infrastructure

The healthcare infrastructure essentially determines both the quantity and quality with which health care services are be delivered in a time of an epidemic. As measures for this determinant we include 4 variables which capture the quantity of hospital beds, nurses and medical practitioners, as well as the quality of the coverage of essential health services. On the one hand, studies report that well structured healthcare resources positively affect a country’s capacity to deal with pandemic emergencies [19–25]. On the other hand, the healthcare infrastructure also greatly impacts the country’s ability to perform testing and reporting when identifying the infected people. In this regard, economies with better structure are able to easily perform mass testing and more detailed reporting [26–28].

#### National health statistics

The physical and mental state of a person play an important role in the degree to which the individual is susceptible to a disease. It is expected that a nation composed of unhealthy individuals should also experience greater consequences of an emergent epidemics [29– 32]. Specifically, metabolic disorders such as diabetes may intensify pandemic complications [33, 34], whereas it has been observed that communicable diseases account for the majority of deaths in complex emergencies [35]. In addition, there is empirical evidence that adequate hygiene greatly reduces the rate of mortality [36, 37]. To quantify the national health characteristics we include 6 variables that asses the level of healthiness among the studied countries.

#### Economic performance

We evaluate the economic performance of a country through 6 variables. This performance often mirrors the country’s ability to intervene in a case of a public health crisis [38–42]. Variables such as GDP per capita have been used in modeling health outcomes, mortality trends, cause-specific mortality estimation and health system performance and finances [43, 44]. For poor countries, economic performance appears to improve health by providing the means to meet essential needs such as food, clean water and shelter, as well access to basic health care services. However, after a country reaches a certain threshold of GDP per capita, few health benefits arise from further economic growth. It has been suggested that this is the reason why, contrary to expectations, the economic downturns during the 20th century were associated with declines in mortality rates [45, 46]. Observations indicate that what drives the health in industrialized countries is not absolute wealth or growth but how the nation’s resources are shared across the population [47]. The more egalitarian income distribution within a rich country is associated with better health of population [48–51]. Nevertheless, it is also known that in better economies the trade interactions and mobility of people is faster, which may enhance the propagation of an transmitted disease [17, 19, 42, 52, 53].

#### Societal characteristics

The characteristics of a society often reveal the way in which people interact, and thus spread the disease. In this aspect, properties such as education and media usage reflect the level of a person’s reaction and promotion of self-induced measures for reducing the spread of the disease [54–58]. Governing behavior such as control of corruption, rule of law or government effectiveness further enhance societal responsibility [59,60]. There are findings which identify the religious view as a critical determinant in the health outcome [61, 62]. Evidently, the religion drives a person’s attitudes towards cooperation, government, legal rules, markets, and thriftiness [63]. Finally, the way we mix in society may effectively control the spread of infectious diseases [17, 53, 64–66]. To measure the characteristics of a society we identify 9 variables.

#### Demographic structure

Similarly to the national health statistics, the demographic structure may evaluate the susceptibility of the population to a disease. Certain age groups may simply have weaker defensive health mechanisms to cope with the stress induced by the disease [67–69]. In addition, the location of living may greatly affect the way in which the disease is spread [70, 71]. To express these phenomena we collect 6 variables.

#### Natural environment

A preserved natural environment ensures healthy lives and promotes well-being for all at all ages. In contrast, countries where natural sustainability is deteriorated and observables such as air pollution are of immense magnitude, are also more vulnerable to epidemic outbreak [6, 7, 72, 73]. However, healthy natural environments also attract a plethora of tourists and thus may help in an easier transformation from epidemic to a pandemic. We gather the data for 4 variables which capture the essence of this socio-economic characteristic [26].

### 2.4 BMA estimation

We use this set of determinants and estimate two distinct BMA models. In the first model the dependent variable is the log of COVID-19 cases per million population, whereas in the second model we investigate the critical determinants of the log of the mortality rate due to the coronavirus. The data gathering and preprocessing procedure is described in Section S1, whereas the mathematical background of BMA together with our inference setup is given in Section S2.2.

Tables 2 and 3 display the respective results. In both tables, the determinants are ordered according to their posterior inclusion probabilities (PIP), given in the second column. PIP quantifies the posterior probability that a given determinant belongs to the “true” linear regression model. Besides this statistic, we also provide the the posterior mean (Post mean) and the posterior standard deviation (Post Std). Post mean is an estimate of the average magnitude of the effect of a determinant, whereas the Post Std evaluates the deviation from this value.

**Table 2:** BMA results with COVID-19 cases per million population as dependent variable.

| Determinant | PIP | Post Mean | Post Std |
| --- | --- | --- | --- |
| Population, total | 0.9783 | −0.3797 | 0.0976 |
| GDP p.c., PPP | 0.6696 | 0.4126 | 0.3021 |
| Gov. health spending p.c., PPP | 0.2898 | 0.1812 | 0.2896 |
| % of population using internet | 0.0285 | −0.0096 | 0.0616 |
| Life expectancy at birth, (years) | 0.0276 | 0.0120 | 0.0785 |
| Mortality rate from lack of hygiene | 0.0234 | −0.0121 | 0.0839 |
| People per sq. km | 0.0194 | 0.0029 | 0.0232 |
| 60%+ catholic population | 0.0146 | 0.0021 | 0.0193 |
| Population age 65+ | 0.0083 | 0.0032 | 0.0414 |
| Physicians p.c. | 0.0067 | 0.0019 | 0.0299 |
| Trade (% of GDP) | 0.0056 | −0.0006 | 0.0111 |
| Nurses and midwives p.c. | 0.0048 | −0.0007 | 0.0136 |
| Number of tourist arrivals | 0.0043 | 0.0004 | 0.0134 |
| Ecological Footprint (gha/person) | 0.0042 | −0.0005 | 0.0184 |
| Yearly passengers carried | 0.0039 | 0.0000 | 0.0111 |
| Rural population (% of total) | 0.0038 | 0.0003 | 0.0084 |
| 60%+ christian population | 0.0037 | −0.0002 | 0.0062 |
| Int. migrant stock (% of population) | 0.0034 | 0.0003 | 0.0093 |
| Yearly avg P.M. 2.5 | 0.0033 | 0.0003 | 0.0091 |
| Government gross debt(% of GDP) | 0.0030 | 0.0001 | 0.0050 |
| Avg. no. of persons in a household | 0.0030 | −0.0002 | 0.0078 |
| Mortality rate from com. diseases | 0.0029 | −0.0002 | 0.0083 |
| Mortality rate from diabetes | 0.0029 | −0.0001 | 0.0063 |
| UHC service coverage index | 0.0028 | −0.0002 | 0.0086 |
| Employment to population ratio (%) | 0.0027 | −0.0001 | 0.0044 |
| GINI index | 0.0027 | 0.0000 | 0.0048 |
| Death rate, crude p.c. | 0.0027 | 0.0000 | 0.0051 |
| Government effectiveness | 0.0027 | 0.0000 | 0.0073 |
| Human capital index | 0.0026 | 0.0001 | 0.0118 |
| Population ages 0-14 | 0.0026 | −0.0002 | 0.0091 |
| Birth rate | 0.0026 | −0.0001 | 0.0086 |
| 60%+ muslim population | 0.0026 | 0.0000 | 0.0043 |
| Rule of law | 0.0026 | 0.0000 | 0.0061 |
| Control of corruption | 0.0025 | 0.0000 | 0.0062 |
| Hospital beds p.c. | 0.0025 | 0.0000 | 0.0058 |

**Table 3:** BMA results with COVID-19 deaths per million population as dependent variable.

| **Determinant** | **PIP** | **Post Mean** | **Post Std** |
| --- | --- | --- | --- |
| GDP p.c., PPP | 0.7797 | 0.5456 | 0.3327 |
| Gov. health spending p.c., PPP | 0.1189 | 0.0735 | 0.2068 |
| Government effectiveness | 0.1032 | −0.0485 | 0.1516 |
| Physicians p.c. | 0.0760 | 0.0436 | 0.1560 |
| Rule of law | 0.0583 | −0.0218 | 0.0940 |
| 60%+ catholic population | 0.0500 | 0.0121 | 0.0566 |
| Population, total | 0.0303 | −0.0068 | 0.0417 |
| Control of corruption | 0.0298 | −0.0102 | 0.0635 |
| Mortality rate from lack of hygiene | 0.0146 | −0.0073 | 0.0650 |
| Population age 65+ | 0.0100 | 0.0048 | 0.0533 |
| Yearly passengers carried | 0.0077 | −0.0014 | 0.0190 |
| Life expectancy at birth, (years) | 0.0061 | 0.0023 | 0.0354 |
| Ecological Footprint (gha/person) | 0.0059 | −0.0012 | 0.0333 |
| People per sq. km | 0.0047 | 0.0006 | 0.0115 |
| Number of tourist arrivals | 0.0046 | −0.0008 | 0.0144 |
| Employment to population ratio (%) | 0.0039 | −0.0005 | 0.0094 |
| % of population using internet | 0.0038 | −0.0002 | 0.0213 |
| Nurses and midwives p.c. | 0.0035 | −0.0007 | 0.0156 |
| Government gross debt(% of GDP) | 0.0029 | 0.0003 | 0.0078 |
| UHC service coverage index | 0.0029 | 0.0007 | 0.0198 |
| Human capital index | 0.0028 | 0.0004 | 0.0189 |
| 60%+ christian population | 0.0027 | −0.0002 | 0.0071 |
| Birth rate | 0.0026 | −0.0005 | 0.0183 |
| Mortality rate from diabetes | 0.0026 | −0.0002 | 0.0089 |
| Mortality rate from com. diseases | 0.0024 | −0.0004 | 0.0145 |
| 60%+ muslim population | 0.0022 | 0.0001 | 0.0058 |
| Avg. no. of persons in a household | 0.0021 | 0.0000 | 0.0085 |
| Int. migrant stock (% of population) | 0.0021 | 0.0001 | 0.0067 |
| Population ages 0-14 | 0.0021 | 0.0000 | 0.0164 |
| Trade (% of GDP) | 0.0019 | 0.0000 | 0.0048 |
| GINI index | 0.0019 | 0.0001 | 0.0055 |
| Yearly avg P.M. 2.5 | 0.0019 | 0.0000 | 0.0071 |
| Hospital beds p.c. | 0.0018 | 0.0000 | 0.0068 |
| Death rate, crude p.c. | 0.0017 | 0.0000 | 0.0043 |
| Rural population (% of total) | 0.0016 | 0.0000 | 0.0049 |

In the estimation procedure we a priori assume that the “true” model consists of the baseline model and 3 additional independent variables. This implies that the prior inclusion probability of each potential determinant is around 0.09. Hence, we can divide the determinants into four groups according to their posterior inclusion probability value [74].

#### Determinants with strong evidence

(PIP > 0.5). The first group describes the determinants which have by far larger posterior inclusion probability than the prior one, and thus there is strong evidence to be included in the true model. We find two variables for which there is such evidence in explaining the coronavirus cases: population size and GDP per capita (p.c.). The population size is negatively related to the number of registered COVID-19 cases per million population, whereas the GDP p.c. exhibits a positive effect on the same variable. In the situation of coronavirus deaths, however, only the GDP p.c. remains a strong predictor, with a positive magnitude.

#### Determinants with medium evidence

(0.5 > PIP > 0.1). One variable displays medium evidence for being a crucial socio-economic determinant of the registered COVID cases 19 – the government health spending, with a positive impact. When looking at the BMA estimation of COVID-19 deaths, besides this determinant, government effectiveness also shows a medium PIP size.

#### Determinants with weak evidence

(0.1 >PIP> 0.01). These are determinants which have a lower posterior than the prior probability to be included in the true model, but still may account for some of the variations in the coronavirus outcome. For the cases per million population there are 5 such determinants, out of which 2 have a negative impact: the percentage of the population using internet and the mortality rate from lack of hygiene. The results suggest that life expectancy, population density and a population consisting of majorly catholic population, have a low evidence for having a negative marginal effect on the observed COVID-19 cases.

There are 7 determinants for which there is low evidence to be included in the true model describing the coronavirus deaths. Four of them have a negative Post Mean: population size, rule of law, control of corruption and mortality rate from lack of hygiene; whereas the presence of catholic religion and the number of physicians pr capita exhibit a positive Post Mean value.

#### Determinants with negligible evidence

(PIP < 0.01). In total, we find negligible evidence for explaining the coronavirus cases in 27 potential determinants and for explaining the coronavirus deaths in 25 potential determinants.

## 3 Discussion

The preliminary analysis suggests that only a handful of socio-economic determinants are able to explain the current extent of the coronavirus pandemic. The sole determinant strongly related to both coronavirus cases and coronavirus deaths is the level of economic development. Wealthier countries display larger susceptibility to the disease. Interestingly, besides the level of economic development, the population size is also a credible predictor of the registered coronavirus cases per million population, with more populated economies showing greater resistance to being infected by the virus.

A plentiful of reasons can be used as a possible interpretation for these results. For instance, it is known that in structured populations, the degree of epidemic spread scales inversely with population size [75]. This is because, everything else considered, in larger populations it is easier to identify and target the critical individuals that are susceptible to the disease [76]. In a similar fashion, various explanations can be found for the observed effect of economic development, such as increased population mobility and aging population. However, it could also be the case that more developed countries have a bigger testing power and provide better evidence for the coronavirus situation. In fact, this may be suggested by our discovery that there is a medium evidence for past government health expenditure to be positively associated with the coronavirus outcome.

Clearly, the interpretation of our analysis requires a more detailed background due to several reasons. Among these reasons is the fact that we include several potential determinants only through crude approximations. In particular, the level of social mixing is given simply as the average number of persons in a household or the dominant religion in the country. We do not follow the exact social network structures within a population. It is evident that the inhomogeneous nature of these spatial patterns has an essential role in propagation of diseases [17]. In this regard, in future versions of this study we aim to incorporate more detailed measures which capture the essence of social connectedness [77] the degree of individualism [78] and/or community mobility data^3^. In addition, the spread of the coronavirus is obviously still in a transient regime. Even though, we include a proxy for the duration of the coronavirus pandemic in each country, this essentially hinders the development of a coherent modeling framework.

These underexpressed effects may play a significant role in the final outcome of the coronavirus pandemic. Nonetheless, in the absence of a unifying framework covering all relevant aspects, our investigation acts as the starting point for the development of a more comprehensive understanding of the socio-economic factors of the coronavirus pandemic. We believe that with the availability of new data and the improved understanding of the dynamics of the coronavirus pandemic, some of these shortcomings will be overcome, yielding a more reliable interpretation of the results.

## Data Availability

References to the raw values of the used data are given in the manuscript. The cleared data is available upon request.

## Supplementary material

### S1 Data description

The data for the dependent variable are taken from Worldometer’s 2019-20 Coronavirus tracker. The tracker offers live coverage of country coronavirus statistics, by collecting data from sources which include Official Websites of Ministries of Health or other Government Institutions and Government authorities’ social media accounts. Because national aggregates often lag behind the regional and local health departments’ data, an important part of the data collection process consists in monitoring thousands of daily reports released by local authorities. The current results were made with data gathered on 11th April 2020.

The data used for calculation of the stringency index and the days since the first regisrered COVID-19 case are gathered from Oxford’s COVID-19 government response tracker. Finally, the data used for measuring the possible socio-economic determinants are gathered from 7 various sources. In particular, the collection is as follows: 25 determinants are from the World Bank’s World Development Indicators (WDI), 3 determinants are respectively from the Nationmaster database (NM) and the World Governance Indicators (WGI), and there is 1 determinant from the International Monetary Fund’s (IMF), the State of Global Air (SGA), the Global footprint network (GFN) and from the United Nations (UN) database. The list of sources together with links to their websites is given in Table S1.

**Table S1:** List of data sources.

| Source | Link |
| --- | --- |
| Covid cases/deaths | www.worldometers.info/coronavirus |
| GFN | data.footprintnetwork.org |
| Gov. Stringency | covidtracker.bsg.ox.ac.uk |
| IMF | www.imf.org/en/data |
| NM | www.nationmaster.com |
| SGA | www.stateofglobalair.org/engage |
| UN | data.un.org |
| WDI | data.worldbank.org/ |
| WGI | info.worldbank.org/governance/wgi |

To reduce the noise from the data we restrict to using only countries with population above 1 million. In addition, we only use countries for which there is data on all of the potential socio-economic determinants. Table S2 gives the countries for which all of these data was available.

**Table S2:** List of countries.

| Country |  |  |  |
| --- | --- | --- | --- |
| Argentina | Finland | Malaysia | Slovakia |
| Australia | France | Mauritius | Slovenia |
| Austria | Germany | Mexico | South Africa |
| Bangladesh | Gyana | Morocco | Spain |
| Belgium | Greece | Myanmar | Switzerland |
| Bolivia | Guatemala | Netherlands | Tanzania |
| Brazil | Honduras | New Zealand | Thailand |
| Bulgaria | Hungary | Nigeria | Turkey |
| Cameroon | India | Norway | United Kingdom |
| Canada | Indonesia | Pakistan | United States |
| Chile | Iraq | Panama | Uganda |
| China | Ireland | Paraguay | Ukraine |
| Columbia | Israel | Peru | Venezuela |
| Costa Rica | Italy | Philippines | Vietnam |
| Croatia | Japan | Poland | Zambia |
| Czech Republic | Jordan | Portugal | Zimbabwe |
| Dominican Rep. | Kazakhstan | Russia |  |
| Ecuador | Kenya | Rwanda |  |
| El Salvador | Madagascar | Serbia |  |

Altogether, we end up with data on 35 variables and 72 countries. Table S3 reports the summary statistics of each variable. We hereby point out that as a measure of the determinant the log of the last observed value is taken, unless otherwise stated in Table S3.

**Table S3:** Summary statistics.

| Variable | Measure | Mean | Std. |
| --- | --- | --- | --- |
| Coronavirus outcome | Coronavirus cases p.m.c. | 4.74 | 2.15 |
|  | Coronavirus deaths p.m.c. | 1.18 | 2.27 |
| Government stringency | Stringency index | −0.31 | 0.88 |
| Epidemic duration | Days since first registered case<sup>a</sup> | 47.14 | 19.12 |
| **Healthcare Infrastructure** |  |  |  |
| Number of physicians | Physicians p.c. | 0.31 | 1.18 |
| Number of nurses | Nurses and midwives p.c. | 1.04 | 1.19 |
| Number of hospital beds | Hospital beds p.c. | 0.84 | 0.85 |
| Health coverage | UHC service coverage index | 4.23 | 0.22 |
| **National health statistics** |  |  |  |
| Birth Rate | Birth rate, crude p.c. | 2.75 | 0.46 |
| Death Rate | Death rate, crude p.c. | 2.02 | 0.32 |
| Life expectancy | Life expectancy at birth, (years) | 4.31 | 0.09 |
| Mortality from diabetes | Mortality rate from diabetes | 2.75 | 0.33 |
| Mortality from communicable diseases | Mortality rate from com. diseases | 2.38 | 0.96 |
| Mortality from hygiene | Mortality rate from lack of hygiene | 0.03 | 2.02 |
| **Economic performance** |  |  |  |
| Economic development | GDP p.c., PPP \$ | 9.74 | 0.96 |
| Labor market | Employment to population ratio (%) | 4.04 | 0.19 |
| Government spending | Gov. health spending p.c., PPP \$<sup>b</sup> | 6.12 | 1.61 |
| Government debt | Government gross debt(% of GDP) | 3.96 | 0.51 |
| Income inequality | GINI index | 3.62 | 0.21 |
| Trade | Trade (% of GDP)<sup>b</sup> | 4.22 | 0.49 |
| **Societal characteristics** |  |  |  |
| Media usage | % of population using internet | 4.02 | 0.51 |
| Education | Human capital index | −0.51 | 0.25 |
| Government effectiveness | Government effectiveness<sup>a</sup> | 0.30 | 0.92 |
| Rule of law | Rule of law<sup>a</sup> | 0.18 | 1.01 |
| Control of corruption | Control of corruption<sup>a</sup> | 0.12 | 1.01 |
| Religion | 60%+ catholic population | 0.40 | 0.49 |
|  | 60%+ christian population | 0.32 | 0.47 |
|  | 60%+ muslim population | 0.12 | 0.33 |
| Household size | Avg. no. of persons in a household | 1.21 | 0.30 |
| **Demographic structure** |  |  |  |
| Elderly population | Population age 65+ (% of total) | 2.21 | 0.72 |
| Young population | Population ages 0-14 (% of total) | 3.13 | 0.37 |
| Population size | Population, total | 17.11 | 1.39 |
| Rural population | Rural population (% of total) | 3.36 | 0.68 |
| Migration | Int. migrant stock (% of population) | 1.05 | 1.62 |
| Population density | People per sq. km | 4.43 | 1.19 |
| **Natural environment** |  |  |  |
| Sustainable development | Ecological Footprint (gha/person) | 1.02 | 0.61 |
| Air Pollution | Yearly avg P.M. 2.5 | 2.95 | 0.62 |
| Air transport | Yearly passengers carried | 8.98 | 2.47 |
| International Tourism | Number of tourist arrivals | 15.66 | 1.41 |

<sup>a</sup> Raw values.
<sup>b</sup> 10 year averages.

## S2 Methods

### S2.1 Stringency index

To calculate our government stringency measure we make use of Oxford’s daily government stringency index. Oxford’s daily government stringency index measures on a scale of 1-100 the variation in daily government responses to COVID-19 by accumulating ordinal data on country social distancing measures on school, workplace and public transport closure; cancellation of public events; restrictions of internal movement; control of international travel and promotion of public campaigns on prevention of coronavirus spread.

To calculate the overall index stringency index *c*_*i*_(*d*_*i*_) at a final date *d*_*i*_ from the provided daily indexes we implement the following procedure. Let *c*_*i*_(*t*) represent the government stringency on day *t*, then our index can be estimated as

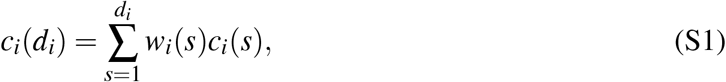

where *w*_*i*_(*s*) are the weights given to each day and *s* = 1 is the day of the first registered case. We use a simple inverse weight procedure by giving larger weights to earlier dates, i.e.,

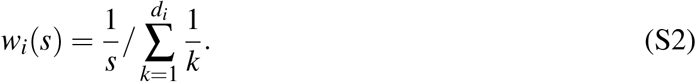

### S2.2 Bayesian model averaging

BMA leverages Bayesian statistics to account for model uncertainty by estimating each possible model, and thus evaluating the posterior distribution of each parameter value and probability that a particular model is the correct one [18]. More precisely, in BMA, the posterior probability for the parameters *g*(*β*_*m*_|*y, M*_*m*_) is calculated using *M*_*m*_ as:

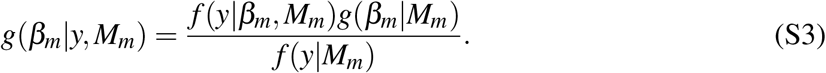

It is clear that the posterior probability is proportional to *f* (*y*|*β*_*m*_, *M*_*m*_), - the likelihood of seeing the data under model *M*_*m*_ with parameters *β*_*m*_, and *g*(*β*_*m*_ | *M*_*m*_) – the prior distribution of the parameters included in the proposed model. By assuming a prior model probability *P*(*M*_*m*_) we can implement the same rule to evaluate the posterior probability that model *M*_*m*_ is the true one, as

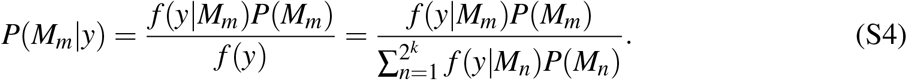

The term *f* (*y* | *M*_*m*_) is called the marginal likelihood of the model and is used to compare different models to each other. The posterior model probability can also be written as

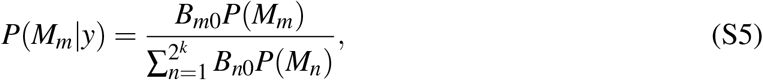

where *B*_*m*0_ is the Bayes information criterion between model *M*_*m*_ and the baseline model *M*_0_. In our case this is the model including government social distancing measures and the length of the coronavirus crisis in the country.

With this setup, we can define the posterior distribution of *β* as a weighted average of the posterior distributions of the parameters under each model using the posterior model probabilities as weights

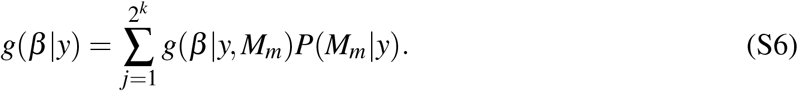

Here, we are interested only in some parameters of the posterior distribution, such as the posterior mean and variance of each parameter. Using equation (S6) we can calculate the posterior mean as:

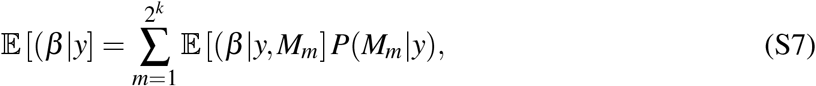

and the posterior variance as:

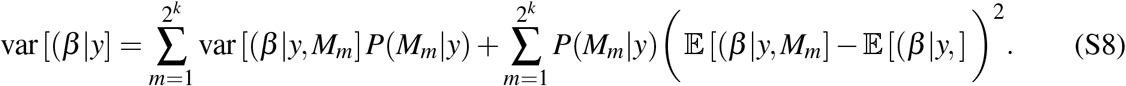

Since the posterior mean is a point estimate of the average marginal contribution, we use it as our measure of the effect of the determinant on the COVID-19 impact.

Another interesting statistic is the posterior inclusion probability *PIP*_*h*_ of a variable *h*, which measures the posterior probability that the variable is included in the ‘true’ model. Mathematically, *PIP*_*h*_ is defined as the sum of the posterior model probabilities for all of the models that include the variable:

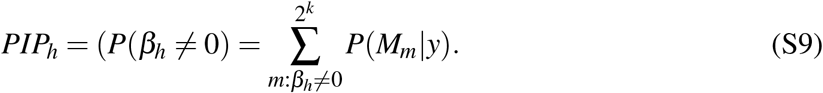

Posterior inclusion probabilities offer a more robust way of determining the effect of a variable in a model, as opposed to using p-values for determining statistical significance of a model coefficient because they incorporate the uncertainty of model selection. According to equations (S3) and (S4), it is clear that we need to specify priors for the parameters of each model and for the model probability itself. To keep the model simple and easily implemented here we use the most often implemented priors. In other words, for the parameter space we elicit a prior on the error variance that is proportional to its inverse, *p*(*σ* ^2^) ≈ 1*/σ* ^2^, and a uniform distribution on the intercept, *p*(*α*) *→*1, while the Zellner’s g-prior is used for the *β*_*m*_ parameters, and for the model space we utilise the Beta-Binomial prior. To estimate the posterior parameters we use a Markov Chain Monte Carlo (MCMC) sampler, and report results from a run with 200 million recorded drawings and after a burn-in of 100 million discarded drawings. The theoretical background behind our setup can be read in Refs. [18, 79–81].

## Footnotes

1 Source: Worldometers coronavirus tracker: https://www.worldometers.info/coronavirus/

2 More about the index developed by the Oxford group can be read at www.bsg.ox.ac.uk/research/publications/variation-government-responses-covid-19

3 Community mobility data is available at https://www.google.com/covid19/mobility/.

